# What interventions are effective and cost-effective for supporting the health and well-being of people with obesity on healthcare waiting lists? A Rapid Review

**DOI:** 10.1101/2024.11.07.24316892

**Authors:** Toby Ayres, Jordan Everitt, Alesha Wale, Chukwudi Okolie, Amy Fox-McNally, Helen Morgan, Hannah Shaw, Jacob Davies, Rhiannon Tudor Edwards, Adrian Edwards, Alison Copper, Ruth Lewis

## Abstract

Currently, there is a significant demand for tier 3 weight management services, with individuals waiting between three and five years to access these services in parts of Wales. This rapid review aimed to identify and synthesise the evidence for the effectiveness of strategies for supporting the health and well-being of individuals with obesity on such waiting lists, with a focus on practical and resource-efficient interventions that can be implemented within current healthcare constraints.

Seven studies were included, and these were published between 2017 and 2024. Studies were conducted in a range of countries, and no relevant UK based study was identified. Studies investigated exercise, physical activity counselling, education and text message-based prehabilitation interventions in people awaiting surgery.

This review did not identify any studies assessing the effectiveness of interventions that could be feasibly implemented or scaled up within the resource limitations of typical tier 3 weight management services in Wales. Most studies required significant resource and input from healthcare professionals, and were delivered in-person at healthcare settings or remotely via teleconferencing. All studies assessed patients with obesity on a waiting list for surgery, but none included a patient population that matched those on waiting lists for tier 3 weight management services in Wales. None of the studies evaluated the cost-effectiveness of interventions.

Overall, we are not confident in the evidence. Most studies were of low quality, with significant methodological and reporting limitations affecting the reliability of their findings. Although we have little confidence in the evidence, there is some evidence from four studies, that suggest exercise interventions may support the quality of life and anthropometric measures of people with obesity waiting for surgery. This evidence could be cautiously considered to inform interventions in practice, but those designing interventions should be mindful of the population and setting in which they are applied. Other interventions, including text message-based prehabilitation interventions, preoperative educational interventions and physical activity counselling interventions were reliant on findings from single low-quality studies. Some of these interventions showed improvements for participant’s quality of life, mental well-being and anthropometric measures.

In relation to obesity weight management services, allocation of resources should allow for conducting and evaluating robust studies and economic evaluations investigating interventions for those awaiting obesity weight management services. Given the current healthcare resource constraints, it may be beneficial to consider the feasibility and scalability of interventions during their design.

## 1. BACKGROUND

### 1.1 Who is this review for?

This rapid review was conducted as part of the Health and Care Research Wales Evidence Centre Work Programme. The question was proposed by Cwm Taf Morgannwg University Health Board Weight Management Service to address the need for feasible, low-resource interventions that could support individuals with obesity on waiting lists for tier 3 services, helping to prevent deterioration in health and well-being during prolonged waiting periods.

### 1.2 Background and purpose of this review

Prevalence of obesity (body mass index [BMI] of ≥30kg/m^2^) has risen rapidly in recent decades and is considered one of the most significant public health challenges worldwide (Watkins, 2024). In Wales, latest data estimates that 25% of adults aged 16 and over are living with obesity (Watkins, 2024). By 2035, it is projected that 11% of the Welsh population will have severe obesity (BMI ≥40kg/m^2^), compared to 8% in England, and 5% in Scotland (Keaver et al, 2020). Currently the UK utilises a four-tier pathway for the management of services to support adults with overweight (BMI 25 – 29.9kg/m^2^) and obesity to lose weight. In Wales specifically, the All-Wales Weight Management Pathway sets out the key elements and principles underpinning the delivery of weight management services. Tier 1 includes brief advice and self-directed materials for overweight adults, Tier 2 includes multicomponent interventions for adults with obesity, Tier 3 includes specialist multi-disciplinary services for adults with severe obesity, and Tier 4 involve specialist surgical services (Welsh Government, 2021).

Obesity prevalence within the Cwm Taf Morgannwg University Health Board area (29% of adults) is higher than the national average (StatsWales, 2023) however, a weight management service operating at tiers 1 to 4 has only been available in the area since February 2023 (Life Sciences Hwb, 2023). Currently, there is a significant demand for tier 3 services, with individuals waiting between three and five years to access these services across parts of Wales (Wales NHS Confederation, 2024). During these prolonged waits, individuals with severe obesity face considerable risk of health deterioration. Due to the high demand on the service leading to extended wait times, and the lack of resources to increase capacity, there is an urgent need for effective interim support strategies for people with severe obesity on waiting lists for tier 3 weight management services in order to prevent their health and well-being deteriorating and promote ‘waiting well’.

Stakeholders highlighted the need for scalable strategies that fit within current resource constraints to support people awaiting tier 3 services to ‘wait well’ and prevent deterioration in health and well-being during this period. Preliminary scoping work conducted for this rapid review identified little secondary or primary research specifically focused on evaluating interventions to support people on waiting lists to attend tier 3 multi-disciplinary services.

Consequently, the focus of this rapid review has been broadened to identify primary research on interventions that could potentially support adults with obesity on any healthcare waiting list.

## 2. RESULTS

### 2.1 Overview of the Evidence Base

The methods and eligibility criteria for the review are presented in Section 5. Eligibility was restricted to comparative primary studies and economic evaluations assessing the effectiveness or cost-effectiveness of interventions aimed at supporting adults with obesity on any waiting list (see section 5.1 for full eligibility criteria). Seven studies were eligible for inclusion. All studies investigated interventions designed to support patients with obesity on surgical waiting lists. None of the studies addressed the stakeholders initial research question which was to support people waiting for tier 3 obesity weight management services.

Two studies were randomised controlled trials (RCTs) while the remaining five studies included two uncontrolled before and after studies, one pilot uncontrolled before and after study, one before and after study that also utilised the comparator usual care group from a previous study, and one non-randomised controlled trial. No economic evaluations were identified. Studies were conducted in a range of countries, including Canada (n=2), Turkey (n=2), Australia (n=1), Germany (n=1), Spain (n=1), and were published between 2017 and 2024. Sample sizes across the studies ranged from six to 305, with six of the seven studies reporting a sample size less than 35. Intervention duration ranged from six-weeks to six-months. Six studies included people with severe obesity (BMI ≥ 40 kg/m^2^ or ≥ 35 kg/m^2^ with comorbidities) who were on a waiting list for bariatric surgery (n=5) or either bariatric or metabolic surgery (n=1). The final study included people with obesity (BMI ≥30 kg/m^2^) on waiting lists for any elective surgery.

Four studies evaluated the effectiveness of interventions, while three focused on assessing their feasibility of implementation. All participants were recruited from surgical waiting lists, either through referral by surgical teams (n=3) or direct contact from researchers (n=1), and three studies did not specify their recruitment methods. The interventions aimed to prepare participants for surgery by targeting improvements in quality of life, physiological, anthropometric, behavioural, functional, or mental health outcomes. In four studies, participants in the intervention group received an intervention in combination with usual care, and outcomes were compared to a control group who also received the usual care component. In these three studies, the usual care component was physical activity counselling (n=2) or nutritional and psychological counselling (n=1). One study compared the effectiveness of a group-based preoperative educational intervention delivered via teleconferencing to an in-person intervention. This study was conducted during the COVID-19 pandemic, when teleconferencing became an alternative to in-person meetings. In the remaining three studies, participants received an intervention, no usual care component was described and outcomes were compared before and after the intervention.

Interventions were based on exercise (n=4), physical activity counselling (n=1), group-based preoperative educational (n=1) and text message-based prehabilitation (n=1). Most interventions were delivered face-to-face or with significant direct healthcare professional input, suggesting a potential mismatch with the need for scalable, low-resource solutions.

Notably, six interventions required direct input from healthcare professionals, with delivery methods including remote teleconferencing (n=2), face-to-face sessions (n=3), and one study that compared both approaches, often conducted in settings like hospitals and university physiotherapy departments. One intervention was delivered through text messaging, where patients received four-weekly text message prompts encouraging healthy behaviours based on Australian physical activity and nutrition guidelines. Exercise interventions frequently required specialist equipment, which, in the case of home-based interventions, was provided to participants along with internet connectivity installation or upgrades where appropriate for use during the study period, adding to the resource demands. Remote interventions conducted via videoconferencing or telehealth required participants to have access to digital devices, internet connectivity, and the necessary skills to use these technologies. Greater detail of the specific interventions being investigated is shown in Table 1.

**Table 1:**
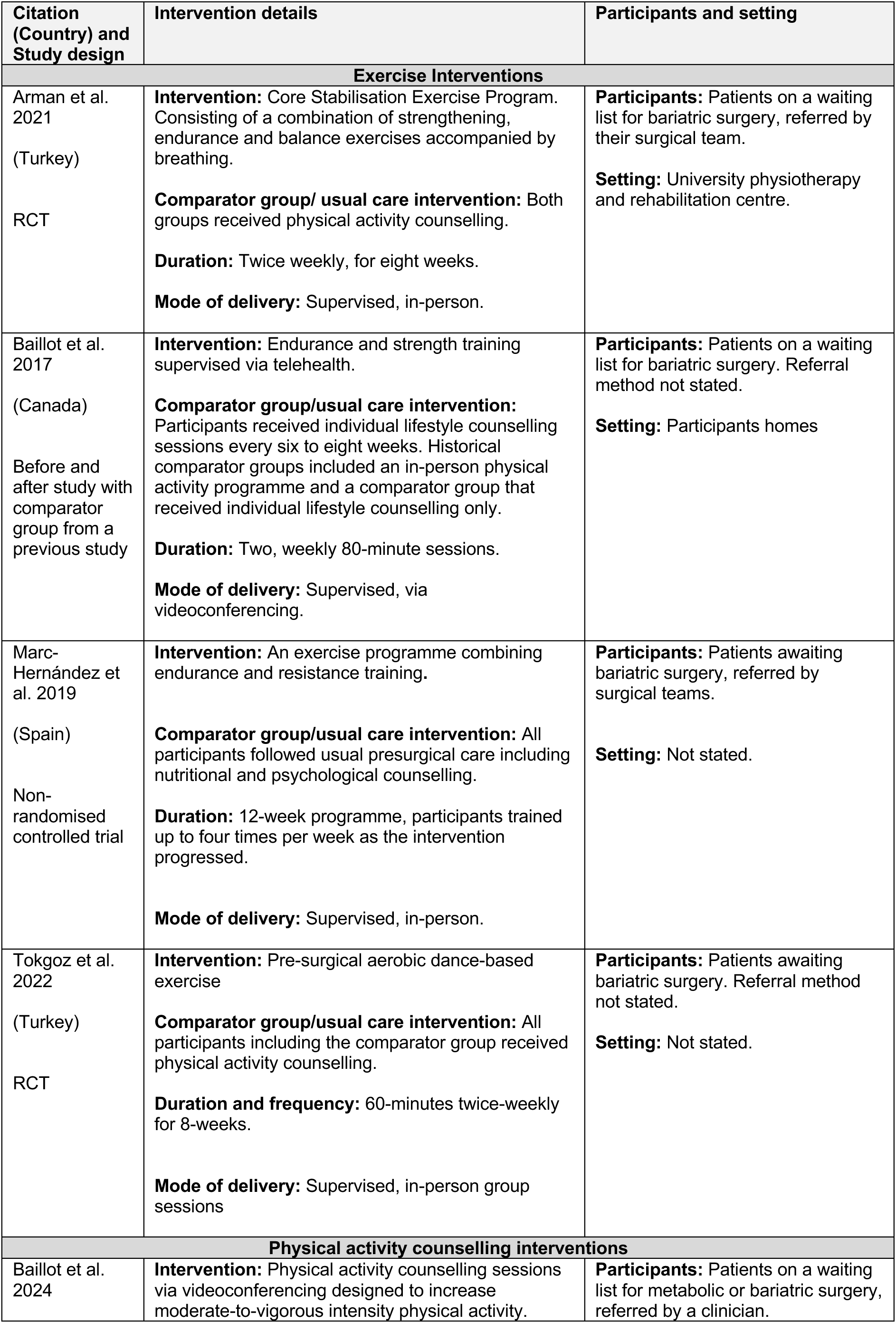
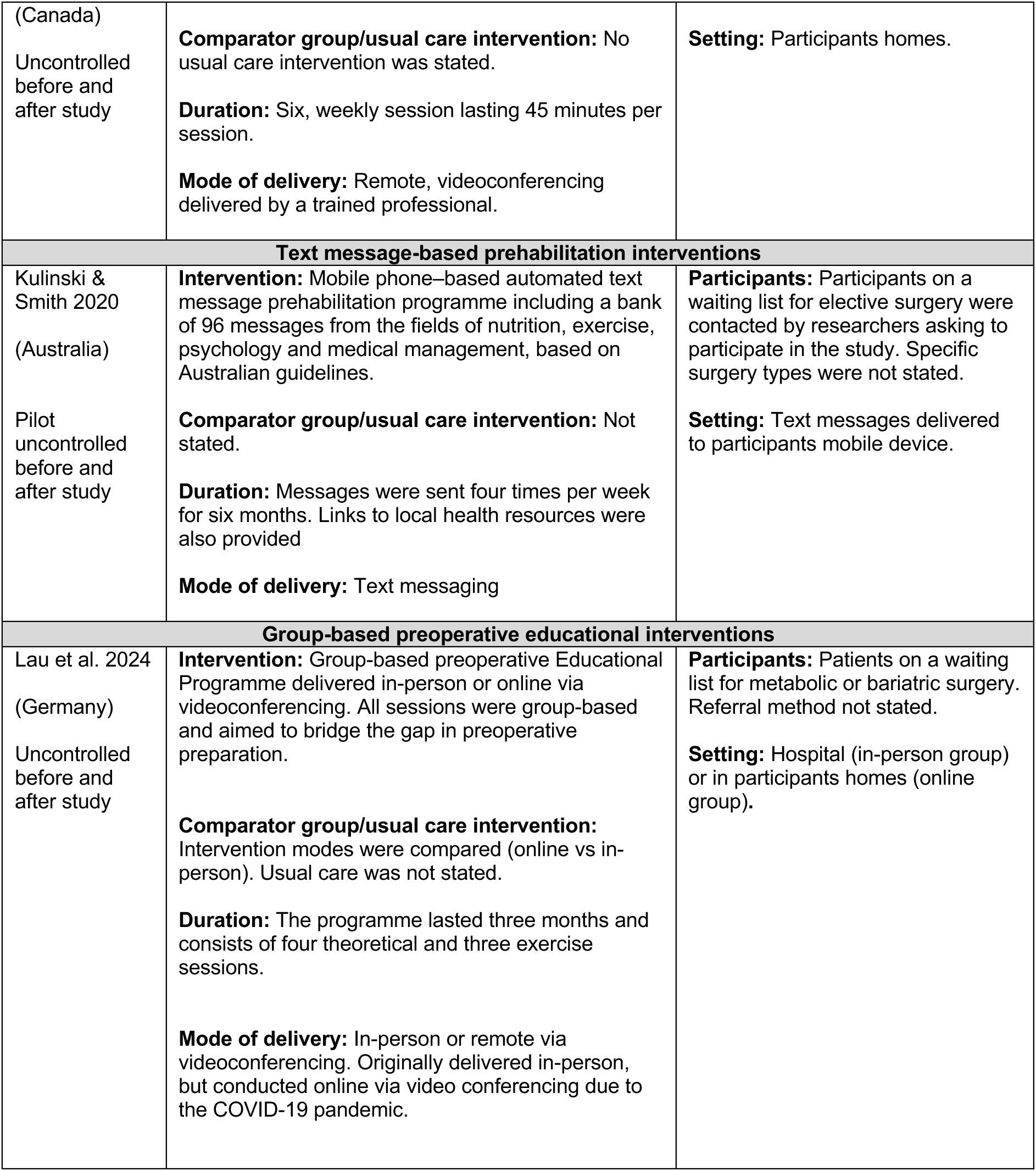
Summary of interventions.

Included studies reported a variety of outcomes that were related to aspects of health and well-being. Commonly reported outcomes included quality of life (n=6) and anthropometric measures such as weight, BMI, fat mass, and fat-free mass (n=5). Mental health measures, including anxiety and depression, were reported in two studies, while fatigue (n=2), stress (n=1), and functional capacity (n=1) were less frequently assessed.

The methodological quality of included studies was assessed in a structured way using established and validated critical appraisal tools for RCTs (Barker et al, 2023) or quasi-experimental studies (Barker et al, 2024). The approach used is reported in more detail in Section 5. All studies had some risk of bias, with quasi-experimental studies rated as low quality (n=5) and RCTs as moderate (n=1) and low (n=1) quality. The moderate quality RCT assessed a Core Stabilisation Exercise Program plus physical activity counselling (usual care), compared with physical activity counselling alone (Arman et al, 2021). A common issue in six studies was the lack of appropriate statistical analysis, including the absence of power calculations and ineffective statistical methods for assessing intervention effectiveness. Three before and after studies did not include control groups and participant outcomes were assessed and comparisons were made before and after the intervention. One RCT lacked blinding, and the other raised concerns about allocation concealment, group similarity at baseline, measurement reliability, and follow-up reporting. Further details of the quality of individual studies are provided in Section 6.3.

A summary of the findings of the effectiveness of each intervention type is reported in the next section. Our certainty (or confidence) in the overall body of evidence on which the findings were based for each outcome measure was assessed according to: the relevance of the available evidence in addressing the review question (directness), the amount and quality of the evidence, the magnitude and direction of effects, similarity of the studies and consistency in the findings (in whether they favour the intervention or control). This information was used to classify the body of evidence for each outcome into one of four levels of certainty: high, moderate, low, and very low. These ratings indicate the degree of confidence we have in the findings, with a high rating indicating, that having assessed the potential problems with the available evidence we are very confident that the summary findings represent the true value of the intervention effect, whilst a very low rating indicates that we have very little confidence that our summary findings represents the true underlying intervention effect. The results of this assessment for the main outcomes, quality of life and mental well-being (anxiety or depression) are provided in Table 2.

**Table 2:**
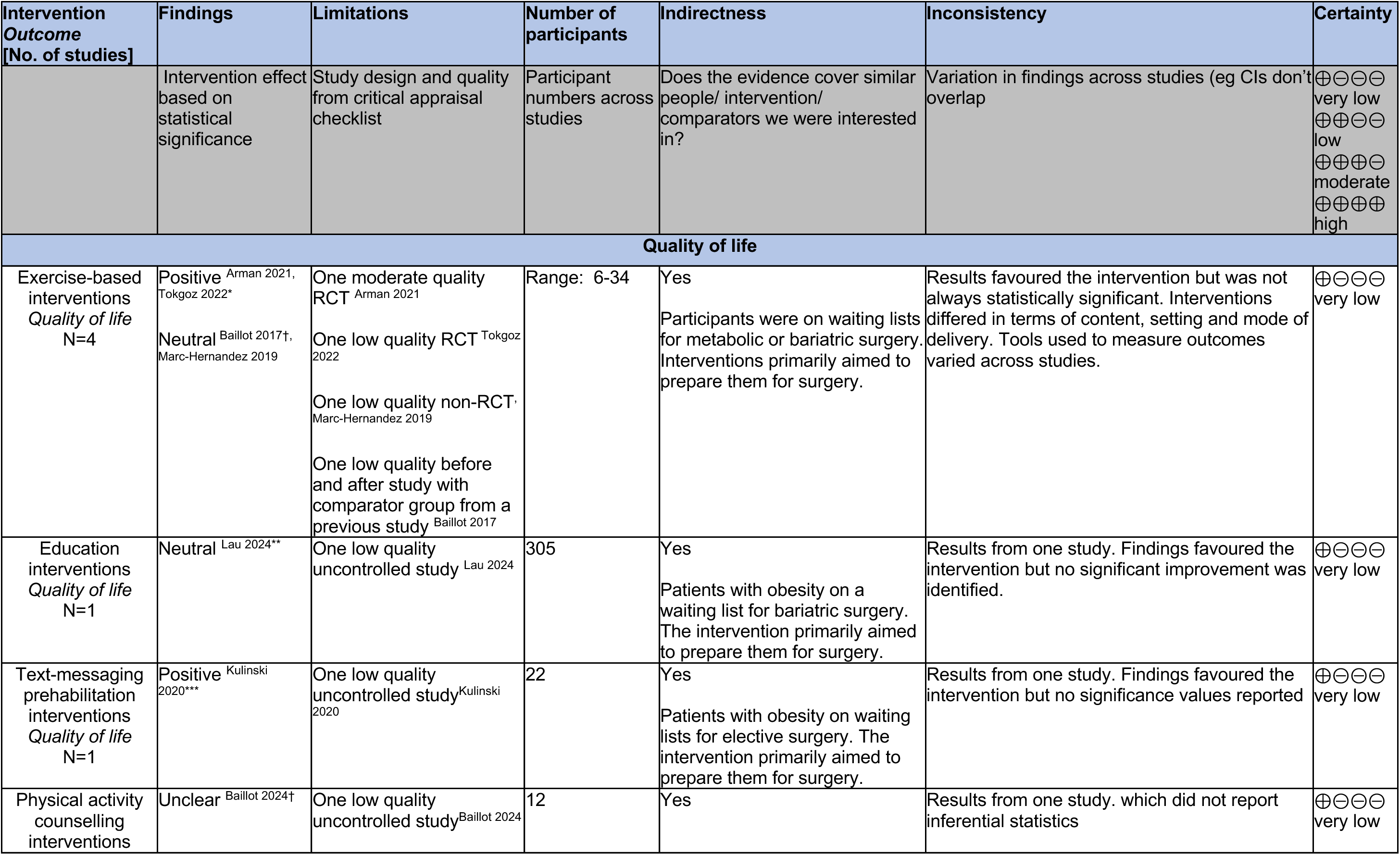
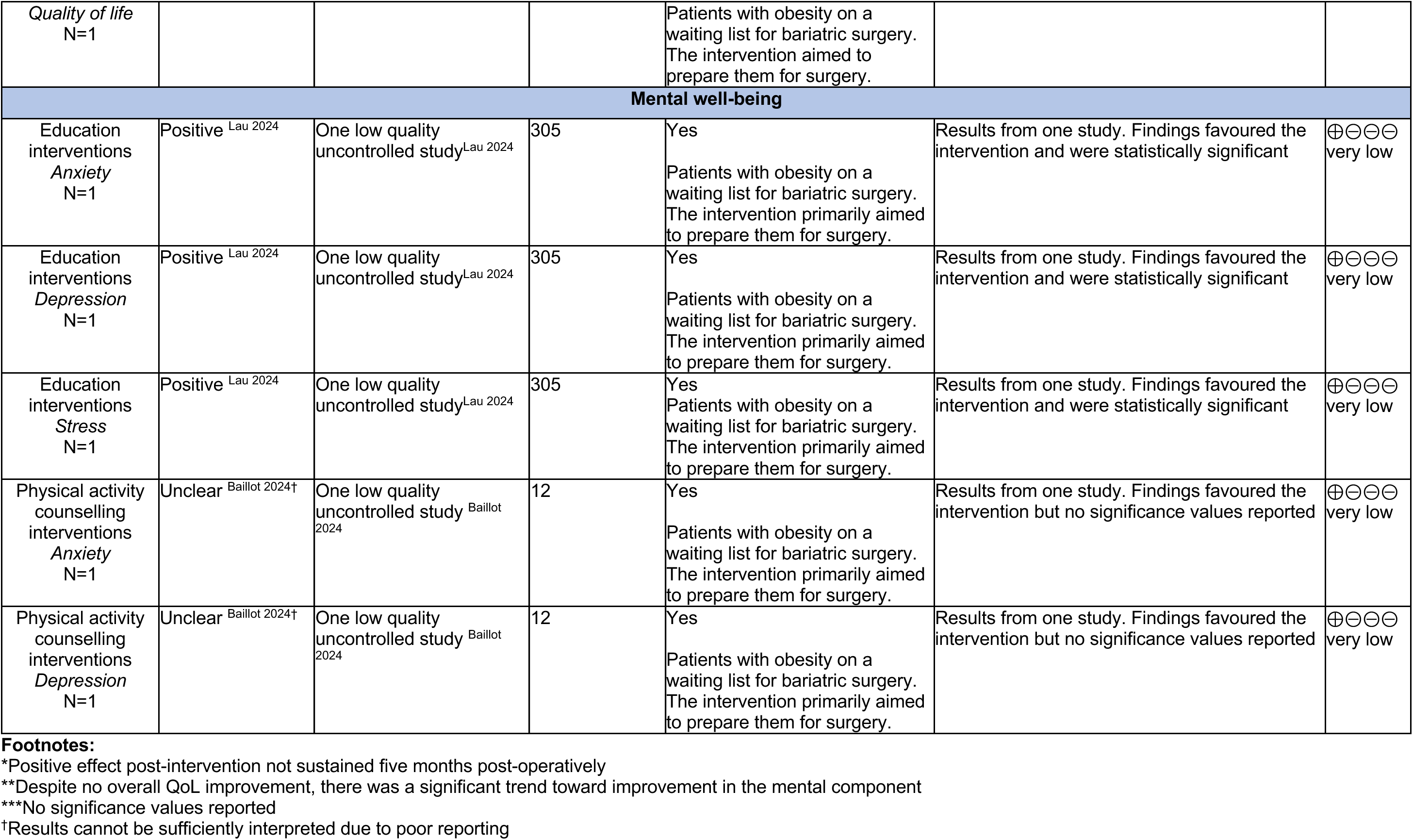
Summary of the evidence.

### 2.2 Effectiveness of exercise interventions

Four studies (Arman et al, 2021; Baillot et al, 2017; Marc-Hernández et al, 2019; Tokgoz et al, 2022) investigated the effects or feasibility of exercise interventions on the health and well-being of patients on waiting lists for bariatric surgery. The types of exercise interventions varied, including combined resistance and endurance programmes (Baillot et al, 2017; Marc-Hernández et al, 2019), a core stability exercise programme (Arman et al, 2021), and aerobic dance-based exercise (Tokgoz et al, 2022). Participants received the exercise intervention in addition to usual care in all four studies. The studies included two RCTs (Arman et al, 2021; Tokgoz et al, 2022), one non-randomised controlled trial (Marc-Hernández et al, 2019) and one before and after study with comparator group from a previous study (Baillot et al, 2017;). Overall, the evidence provided by these studies was of very low certainty.

#### Quality of life outcomes

- Arman et al (2021) assessed the impact of a supervised core stability exercise programme in people with obesity awaiting bariatric surgery. Both the intervention group and control group received physical activity counselling as part of usual care. After the eight-week intervention, a statistically significant difference in Obesity Specific Quality of Life Questionnaire (OSQOL) scores were observed in both groups and between-groups (p=0.001). The intervention significantly improved their OSQOL scores (ES= 0.41, p=0.005), whereas the control group significantly worsened (ES= −0.48, p=0.011).
- Two studies explored the effectiveness of supervised resistance and endurance exercise interventions in people with obesity awaiting bariatric surgery (Baillot et al, 2017; Marc-Hernández et al, 2019). Baillot et al (2017) assessed the feasibility of implementing a home-based videoconferencing intervention and found no significant difference in weight-related quality of life (WRQOL) scores compared to usual care (p≥0.4) or an in-person intervention (p=0.9) group from a previous study (descriptive statistics to indicate the direction of effect were not reported). Marc-Hernández et al (2019) used the 36-Item Short Form Survey (SF-36) questionnaire to assess the impact of a supervised in-person intervention. While the intervention group increased mean scores in all domains, significant between-group differences in favour of the intervention group were reported for only two of eight scales which included: physical functioning (p=0.026), and general health perception (p=0.005), a significant difference was apparent for the physical component summary (p=0.046) as well.
- A group-based aerobic dance intervention (Tokgoz et al, 2022) significantly improved Impact of Weight on Quality of Life-Lite (IWQOL-Lite) scores in people with morbid obesity awaiting bariatric surgery. The intervention group improved their scores immediately post-intervention (p>0.005), whereas control group scores significantly worsened following eight-weeks of physical activity counselling. The improvement in scores shown in the intervention group were greater than that of the control group post-intervention (mean difference: −14.46 ± 22.08 vs 1.64 ± 22.08, p=0.007).

#### Anthropometric outcomes

- Arman et al (2021) measured the effect of the intervention on a range of body composition measures including body weight, BMI, fat mass and fat free mass. No significant differences were reported for either group following the programme (p > 0.05).
- Baillot et al (2017) found no significant differences in BMI or fat mass in participants undergoing the videoconferencing intervention compared to the usual care or in-person programme (p>0.2).
- Tokgoz et al (2022) reported no significant between-group differences for body weight, BMI or fat mass post-intervention (p>0.05).
- Results reported by Marc-Hernández et al (2019) showed the intervention group obtained significant reductions in all anthropometric and body composition variables evaluated total weight (−7.3 ± 4.1 kg, p<0.01), BMI (− 2.6 ± 1.5, p=0.001), fat mass (−7.1 ± 4.7 kg, p <0.01), and waist circumference (−5.3 ± 2.1 cm, p<0.01) except in the fat-free mass (kg), which remained constant. In addition, all variables excluding fat-free mass (kg), showed significant reductions that were superior to the control group.

#### Functional outcomes

- Arman et al (2021) found that the intervention group (IG) significantly improved their 6-Min Walk Test (MWT)-distance values after eight weeks (MD= 146 ± 42.33 [95% CI: 115.71 to 176.28]). The control group 6-MWT-distance values significantly worsened after eight-weeks (MD= −36.63 ± 50.91 [95% CI:-70.83 to −2.43]). There was a statistically significant difference in 6 MWT-distance scores between the groups after the programme in favour of the exercise group (p<0.05). Cohen’s d indicated on all scores of 6 MWT-distance ranged from 0.03 to - 0.78 for the exercise group and - 0.05 to 0.26 for the control group.
- Tokgoz et al (2022) measured fatigue using the Fatigue Severity Scale (FSS), and reported significant improvements from baseline to post intervention. These improvements were significantly greater than the control group (MD= −2.89 ± −1.13 vs −0.49 ± 1.26, p=0.001).

#### Bottom line results for exercise interventions

Overall, our certainty in the findings that exercise interventions are effective in improving the quality of life of people with obesity awaiting surgery is very low (Table 2). The evidence base was limited, with only four relevant studies identified three of which were rated as low methodological quality and one study assessed the feasibility of implementing their intervention. The one moderate quality RCT reported statistical improvements in quality of life following exercise interventions. However, the lack of blinding of participants, outcome assessors and those delivering the treatment limited the strength of these findings. The differences in study design, intervention characteristics such as exercise type, and mode of delivery among these studies limit our confidence in the findings. Studies also used a variety of measurement tools to assess quality of life outcomes, which complicates comparisons and further limits certainty. Evidence for the impact of exercise on anthropometric measures, such as body weight and BMI varied, with a majority of studies showing no significant effect of the intervention. There is also very little evidence supporting improvements in functional outcomes.

### 2.3 Effectiveness of text message-based prehabilitation interventions

One study (Kulinski and Smith 2020) examined the feasibility of implementing of a text message-based prehabilitation intervention on patients with obesity awaiting elective surgery. This study was a before and after pilot study conducted in Australia. This study was judged to be of low quality and only provided descriptive analysis of outcomes relevant to this rapid review.

#### Quality of life outcomes

- Kulinski and Smith (2020) found that 61% (n=11) of participants reported that their overall health score, as indicated by the quality of life EQ-5D-3L visual analogue scale improved after completing the programme. Twenty-seven percent (n=5) of participants reported improved general health-related quality of life.

#### Anthropometric outcomes

- Kulinski and Smith (2020) reported that 40% (n=8) of participants lost at least 2 kg over the course of the intervention. Those who did lose weight lost a mean of 6 kg over six months (range 2– 12 kg), with a mean weight loss of 1.7 kg for the entire cohort.

#### Bottom line results for text message-based prehabilitation interventions

The evidence for the effectiveness of text message-based prehabilitation interventions for improving quality of life, weight is of very low certainty, is reliant on a single low-quality study that lacked appropriate statistical analysis and reporting. Due to the significant methodological limitations and reliance on a single feasibility study, the effectiveness of this intervention remains unclear.

### 2.4 Effectiveness of group-based preoperative educational interventions

One study (Lau et al., 2024) examined the impact of a group-based educational intervention on mental health outcomes, comparing videoconferencing and face-to-face delivery modes of delivery. This low quality, before and after study conducted in Germany provided very low certainty evidence for the effectiveness of group-based preoperative educational interventions.

#### Mental well-being outcomes

- Anxiety and depression scores, measured using the GAD-7 and PHQ-9 questionnaires respectively, were significantly improved following completion of the study (anxiety: –1.1 units ± 4.6, z = −3.914, p<0.001; depression: –0.9 units ± 4.6, z = –3.771, p<0.001). Participants also significantly reduced perceived stress scores (PSQ-20), overall stress reduced between by 4.6 ± 15.6 points (z = –4.976, p<0.001). In particular, the subscale “worries” showed an average improvement of –7.9 ± 22.7 points (z = –5.761, p<0.001). A mean reduction of five points was achieved for the subscale “tension” (z = –4.237, p<0.001) and for the subscale “demands” (z = −4.944, p<0.001). There were no significant changes on the “joy” subscale. There were no interaction effects between the two subgroups indicating that the mode of delivery (face-to-face vs. videoconference) did not significantly influence outcomes.

#### Quality of life outcomes

- There was no significant change in the physical component of the SF-12 however, there was a trend toward improvement in the mental component of quality of life, with a mean increase of 2.3 points ± 10.8 (z = 3.022, p=0.003).

#### Anthropometric outcomes

- No significant differences in weight or BMI were reported following completion of the intervention (weight: −0.3 kg ± 8.7, z = −0.561, p=0.575; BMI: −0.1 units, ± 3.1, z= - 0.277, p=0.782).

#### Bottom line results for group-based preoperative educational interventions

The evidence for group-based educational interventions, comparing videoconferencing and face-to-face delivery modes, is of very low certainty, with findings from a single low-quality study. Results for both interventions indicate improvements in mental well-being outcomes, including anxiety, depression, and stress reduction. However, there were no significant changes in quality of life, with only a trend towards improvement in the mental component, and no significant changes in the physical component. The interventions showed no effect on anthropometric outcomes, such as weight or BMI. The mode of delivery (face-to-face vs. videoconference) did not significantly influence the results.

### 2.5 Effectiveness of physical activity counselling interventions

One study (Baillot et al., 2024) evaluated the feasibility, acceptability, and effectiveness of a telehealth-based physical activity counselling intervention designed to increase moderate-to-vigorous physical activity in adults awaiting metabolic or bariatric surgery. This low quality, before and after study was conducted in Canada. The study’s limited statistical analysis undermines the precision and clarity of the findings, making the effectiveness of the intervention unclear.

#### Mental well-being and quality of life outcomes

Baillot et al (2024) found at least one minimally important difference (improvement in RAND-36 score > 25% post-intervention) for psychosocial outcomes (anxiety and depressive symptoms, quality of life) was reached by 90.9% (n=10) of participants during and after the intervention. This low-quality study does not report inferential statistics for these outcomes; therefore, the statistical significance is unclear.

#### Bottom line results for physical activity counselling interventions

The evidence for the effectiveness of physical activity counselling interventions is of very low certainty and is limited to a single low quality feasibility study. While a high proportion of participants showed improvements in some mental well-being and quality of life domains, the poor reporting restricts the ability to gauge the true effectiveness of the intervention. The results suggest potential benefits in improving psychosocial outcomes, but the overall evidence is very low certainty and inconclusive.

## 3. DISCUSSION

### 3.1 Summary of the findings

This rapid review aimed to identify and synthesise the evidence for the effectiveness of interventions to support the health and well-being of people with obesity on waiting lists, with a focus on identifying practical and resource-efficient interventions that can be implemented within the current healthcare constraints of tier 3 weight management services in Wales.

Seven primary studies were identified. These assessed the effectiveness or feasibility of implementing exercise, physical activity counselling, education or text messaging-based interventions. The findings of these studies indicated that these interventions can improve quality of life, anxiety, stress and depressive symptoms, as well as anthropometric measures (body weight, BMI, fat mass, fat-free mass) of patients awaiting surgery. However, the evidence supporting these findings was deemed to be of very low certainty.

Four studies investigating exercise interventions were identified, while the remaining intervention types were reliant on findings from a single study. Although exercise interventions demonstrated positive results for improving quality of life, interventions varied greatly in terms of 1) exercise type (e.g, a Core Stability Exercise Programme and an aerobic based dance intervention), 2) mode of delivery (e.g, supervised in-person or remote delivery via teleconferencing), 3) study design and aim (e.g, RCTs, non-RCTs and uncontrolled studies, and one study assessed the feasibility of implementing an intervention), and 4) quality of life outcomes and tools used to assess them. This, as well as the studies being predominantly of low methodological quality, means that we are not confident in the evidence.

The stakeholders of this rapid review aimed to use findings to inform support strategies for people with obesity on waiting lists for tier 3 weight management services. However, it is likely that the type of interventions assessed in the included studies would not be feasible for implementation within this context. The interventions were resource-intensive and most were delivered in-person, which contrasts with our stakeholder need for brief and low-resource solutions that are scalable to a large number of patients waiting for treatment. One study (Kulinski and Smith 2020) assessed an automated text message-based prehabilitation intervention, which may be better suited to weight management service waiting list constraints; however, this study assessed feasibility of implementation, not effectiveness.

### 3.2 Strengths and limitations of the available evidence

The evidence included in this rapid review was impacted by significant methodological limitations. Most studies were rated as low quality due to not including a control group, absence of blinding in RCTs, and inadequate statistical analysis to support the validity of the findings. These issues contributed to a very low certainty of evidence, which makes it challenging to draw robust conclusions about the effectiveness of the interventions evaluated. Furthermore, three studies assessed the feasibility of an intervention, so effectiveness could not be determined.

Several key evidence gaps further limit the applicability of the findings to the stakeholder context. Firstly, there were no UK-based studies included in the review, and the geographic scope of the studies may affect the generalisability of the results, particularly given differences in healthcare systems, patient demographics, and service availability.

Additionally, most of the included studies focused on individuals on waiting lists for bariatric or metabolic surgery, which does not fully align with a population that is of direct relevance to the stakeholders i.e. people with obesity waiting for tier 3 services. In the UK, patients undergoing bariatric surgery are typically at tier 4 in the weight management pathway and have usually received tier 3 treatment prior to referral for surgery. Therefore, these patients are less representative of the population of primary interest to stakeholders. Additionally, only one study specifically targeted the stabilisation and improvement of mental health during the waiting period, revealing a significant gap in evidence.

We didn’t identify any relevant economic evaluations and no study included an assessment of the cost-effectiveness of interventions, an essential factor for determining the practicality of implementing these interventions within constrained healthcare budgets.

The review did not find any studies that could be feasibly and scalably implemented within current healthcare constraints faced by obesity weight management services. The reliance of technologies (e.g, laptops for videoconferencing) in some interventions also raises equity concerns, as digitally excluding participants—particularly those without reliable internet access, digital devices, or the necessary digital literacy—could disproportionately impact vulnerable groups who are already at a disadvantage in accessing healthcare services. These barriers could limit the reach and effectiveness of such interventions, undermining their potential impact.

### 3.3 Strengths and limitations of this Rapid Review

The methods used for this rapid review were based on an abbreviated systematic review approach, following established guidelines in an attempt to capture all relevant publications with minimal risk of bias in a timely manner. Included studies were systematically identified through an extensive search of electronic databases and grey literature.

However, the rapid review faced notable limitations. During preliminary work, no evidence directly addressing the stakeholders’ original question was identified. As a result, the review’s scope was broadened to capture a wider range of studies. This adjustment, while necessary to ensure potentially relevant research was identified, led to the inclusion of studies of interventions that did not directly address the initial research question. The broadened scope also resulted in the inclusion of studies that could not be feasibly or scalably implemented within current healthcare constraints experienced by stakeholders and were not specifically focused on the population of interest—individuals with obesity on waiting lists for tier 3 weight management services.

Additionally, poor reporting of results in several studies made it challenging to interpret findings accurately. Despite efforts to ensure a robust synthesis and consistency checks at each stage of the review, these reporting issues affected the overall clarity and applicability of the evidence.

### 3.4 Implications for policy and practice

Although we have little confidence in the evidence, there is more evidence of better quality (including two RCTs and two controlled studies) that suggest exercise interventions may support the quality of life of people with obesity waiting for surgery, with no studies reporting a decline in quality of life. This evidence could be cautiously considered to inform interventions in practice but those designing interventions should be mindful of the population and setting in which they are applied.

The findings of this rapid review highlight the need for increased resource allocation to conduct and rigorously evaluate robust studies and economic evaluations investigating waiting list interventions for individuals with obesity. A particular focus should be on UK-based interventions for those awaiting weight management services. When designing interventions decision makers should consider both the feasibility to implement and scalability within the current healthcare resource constraints.

Emerging approaches in the UK show a shift towards digital health solutions to improve the availability of weight management support. This may provide an alternative option for those on waiting lists as well as people in areas where multidisciplinary weight management services are not yet provided. The National Institute for Health Excellence (NICE, 2023) Health Technology Evaluation (HTE14) of digital technologies highlights the potential of applications to deliver multidisciplinary weight-management services and prescribe and/ or monitor weight-management medicine for adults with obesity. These digital platforms could support increased capacity, or reduced demand on clinician time as well as cutting wait times. When considering digital solutions, NICE (HTE14) stresses the importance of evaluating these digital interventions to ensure they are safe, effective, and accessible. While some patients may prefer to access services remotely via digital technologies, it is imperative that services also consider and address the risk of digital exclusion, particularly for those who may need additional support. For example, those at greatest risk of digital exclusion may suffer from a visual, hearing or cognitive impairment, reduced manual dexterity, a learning disability or being unable to read English (NICE, 2023). Furthermore, considering the impact of a shift to digital solutions on those with limited access to technology or low digital literacy is crucial to prevent widening health inequalities.

### 3.5 Implications for future research

Future research should prioritise the development and evaluation of brief, scalable interventions tailored to low-resource settings, specifically designed to support patients to ‘wait well’ without further health deterioration while on waiting lists for tier 3 services. The current evidence base lacks practical solutions that directly address the resource constraints faced by weight management services, highlighting the need for studies that develop and test interventions which are both effective and feasible within these contexts.

Research efforts should focus on exploring digital health solutions, remote support, and minimal contact models that minimise the need for specialist equipment and extensive healthcare professional input and promote equity, including considerations for those at risk of digital exclusion. These approaches could provide scalable options that better meet the needs of healthcare providers and patients in low-resource settings. Additionally, future research should focus on interventions that address the specific needs of individuals on waiting lists for tier 3 weight management services, emphasising the development of interventions that balance effectiveness with feasibility and accessibility. It is crucial that future studies undertake rigorous evaluations, including cost-effectiveness analyses, to determine the clinical and financial viability of interventions. Understanding the economic impact of implementing these interventions will help stakeholders make informed decisions about adopting these solutions.

### 3.6 Economic considerations*

- There is a lack economic evidence on the impact of implementing interventions for supporting the health and well-being of people with obesity on healthcare waiting lists.
- Obesity is a considerable and prevalent public health issue in Wales that incurs significant economic cost. Obesity costs NHS Wales £73 million per annum (Welsh Government, 2011). If rates of overweight and obesity continue to rise in line with recent trends, by 2050, they will cost NHS Wales £465 million per year, with a cost to the wider society and economy of £2.4 billion (Public Health Wales, 2016).

Obesity and overweight can impact one’s ability to enter and remain in work. A press release by the Health Secretary from October 2024 suggested illnesses caused by obesity cause people to take an extra four sick days a year on average (The Telegraph, 2024). Inability to access management services and support while awaiting services may contribute to such absenteeism.

## Data Availability

All data produced in the present study are available upon reasonable request to the authors

## Abbreviations

BMI: Body Mass Index
CI: Confidence interval
COVID-19: Coronavirus disease
CSEP: core stabilisation exercise program
EG: Exercise group
ES: Effect size
EQ-5D-3L: EuroQol-5 dimension 3-level questionnaire
EWL: Excess weight loss
FACIT-F: Functional Assessment of Chronic Illness Therapy – Fatigue
FSS: Fatigue Severity Scale
GAD-7: Generalized Anxiety Disorder Questionnaire - 7
GRADE: Grading of Recommendations, Assessment, Development, and Evaluations
HTE: Health technology evaluation
ICTRP: International Clinical Trials Registry Platform
IWQOL-Lite: Impact of Weight on Quality of Life-Lite
JBI: Johanna Briggs Institute
Kg: Kilograms
MD: Mean difference
MWT: Minute walk test
MVPA: moderate-to-vigorous physical activity
NICE: National Institute for Health and Care Excellence
OECD: Organisation for Economic Co-operation and Development
OSQOL: Obesity Specific Quality of Life
PAC: physical activity counselling
PHQ-9: Patient Health Questionnaire - 9
PICO: Population, intervention, comparator, outcome
PMOABS: people with morbid obesity awaiting bariatric surgery
PRISMA: Preferred Reporting Items for Systematic reviews and Meta-Analyses
PSADBE: Pre-surgical aerobic dance-based exercise
PSQ-20: Perceived Stress Questionnaire - 20
PreSET: Pre-Surgical Exercise Training intervention
QoL: Quality of Life
RCT: Randomised controlled trial
SD: Standard deviation
SF-12: 12-Item Short Form Survey
SF-36: 36-Item Short Form Survey
TelePreSET: Telehealth Pre-Surgical Exercise Training intervention
TELE-BariACTIV: TELEhealth BARIatric behavioural intervention
UK: United Kingdom
WRQOL: weight-related quality of life

## RAPID REVIEW METHODS

### 5.1 Eligibility criteria

We searched for primary sources to answer the review question: what interventions are effective and cost-effective for supporting the health and well-being of people with obesity on healthcare waiting lists?

**Table 3:**
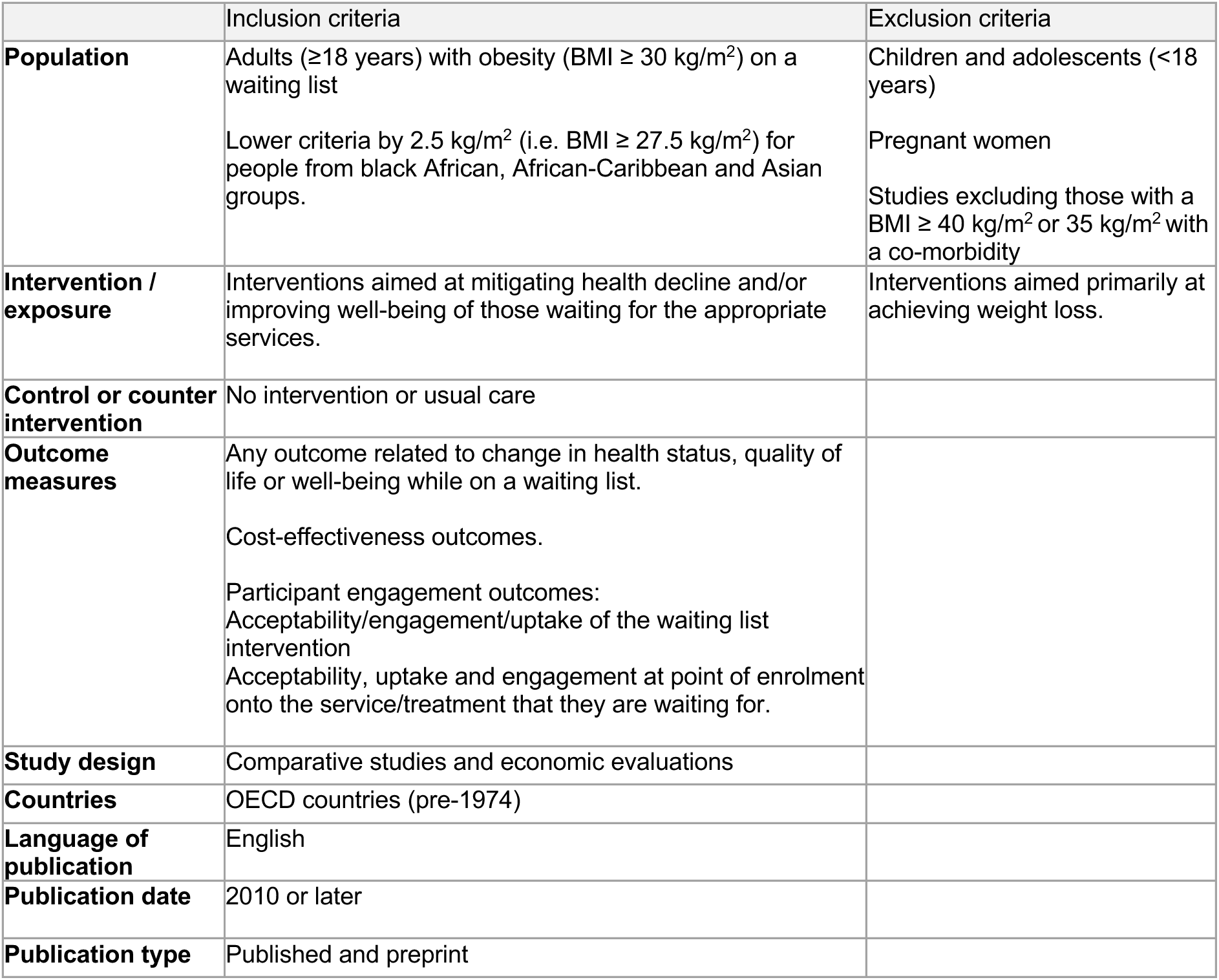
Eligibility Criteria.

|  | Inclusion criteria | Exclusion criteria |
| --- | --- | --- |
| **Population** | Adults ($\geq 18$ years) with obesity ($\text{BMI} \geq 30 \text{ kg/m}^2$) on a waiting list<br>Lower criteria by $2.5 \text{ kg/m}^2$ (i.e. $\text{BMI} \geq 27.5 \text{ kg/m}^2$) for people from black African, African-Caribbean and Asian groups. | Children and adolescents ($< 18$ years)<br>Pregnant women<br>Studies excluding those with a $\text{BMI} \geq 40 \text{ kg/m}^2$ or $35 \text{ kg/m}^2$ with a co-morbidity |
| **Intervention / exposure** | Interventions aimed at mitigating health decline and/or improving well-being of those waiting for the appropriate services. | Interventions aimed primarily at achieving weight loss. |
| **Control or counter intervention** | No intervention or usual care |  |
| **Outcome measures** | Any outcome related to change in health status, quality of life or well-being while on a waiting list.<br>Cost-effectiveness outcomes.<br>Participant engagement outcomes:<br>Acceptability/engagement/uptake of the waiting list intervention<br>Acceptability, uptake and engagement at point of enrolment onto the service/treatment that they are waiting for. |  |
| **Study design** | Comparative studies and economic evaluations |  |
| **Countries** | OECD countries (pre-1974) |  |
| **Language of publication** | English |  |
| **Publication date** | 2010 or later |  |
| **Publication type** | Published and preprint |  |

### 5.2 Literature search

A search was conducted of the electronic bibliographic databases: Medline, Embase, PsycINFO, and CINAHL Plus with Full Text. The search strategy used for Medline is available in appendix 1. A search for ongoing and completed trials was conducted in Clinicaltrials.gov, and WHO International Clinical Trials Registry Platform (ICTRP). Citation tracking from sources identified during the preliminary stages was also undertaken, as well searching reference lists of potentially relevant reports previously conducted by the review team. All searches were conducted between 22/07/2024 and 23/07/2024. Terms used to cover the population and intervention elements of the eligibility criteria included those to describe obesity and phrase variations to describe waiting lists or waiting for support. Searches were limited to studies published after 2010, the year that the All-Wales Obesity Pathway was introduced, and to those published in the English language.

### 5.3 Reference management

All citations retrieved from the database searches were imported or entered manually into EndNote and duplicates removed by a single reviewer. The citations that remained were exported and imported to the systematic reviewing platform Rayyan for study selection.

### 5.4 Study selection process

All titles and abstract were screened independently in duplicate with disagreements settled by discussion and consensus within the review team. This process was repeated for screening records at full-text.

### 5.5 Data extraction

Data extracted was conducted by a single reviewer and was consistency checked by a second reviewer. Information extracted includes:

- Citation Study design
- Intervention
- Comparator
- Study aim
- Data collection methods and dates
- Outcomes reported
- Sample size
- Participants
- Setting
- Key findings
- Observations/Notes

### 5.6 Study design classification

The included studies were classified as quasi-experimental studies or RCTs. No economic evaluations were included following the study selection process.

### 5.7 Quality appraisal

The JBI critical appraisal checklists for quasi-experimental studies (Barker et al, 2024) and RCTs (Barker et al, 2023) were used to assess the methodological quality of each included study. These checklists are not designed to assign an overall score to each study.

Quality assessment was undertaken in duplicate by two independent reviewers. Any discrepancies were discussed and resolved between reviewers. The quality assessment of individual studies can be seen in section 6.3.

### 5.8 Synthesis

Data was synthesised narratively to provide a collective interpretation of the evidence.

### 5.9 Assessment of body of evidence

An assessment of the overall body of evidence was made based on the relevance of the available evidence in addressing the review question, the amount and quality of the evidence, the magnitude and direction of effects, and consistency in the findings. This information is provided for the main outcomes (quality of life and mental well-being) in Table 2, and was used to classify the body of evidence for each outcome as: Very low certainty; Low certainty; Moderate certainty; and High certainty.

## EVIDENCE

### 6.1 Search results and study selection

A visual representation of the flow of studies throughout the review can be found in Figure 1. A total of 3,299 records were retrieved via the searches and, 1,953 records remained following deduplication. A total of 24 articles were screened at full text..

Seven studies were included in the rapid review. Of which, two studies were RCTs, and five were quasi-experimental, including one non-randomised controlled trial, a before and after study that used comparator groups from a previous study, two uncontrolled before and after studies, and one uncontrolled pilot before and after study.

In order to identify studies relevant to the context of weight management services waiting lists, intervention type will be categorised and method of delivery will be described. Relevant delivery methods include but will not be limited to:

- Digital
- Remote
- Self-directed
- Telephone
- Letters/postal
- Brief interventions

**Figure 1.**
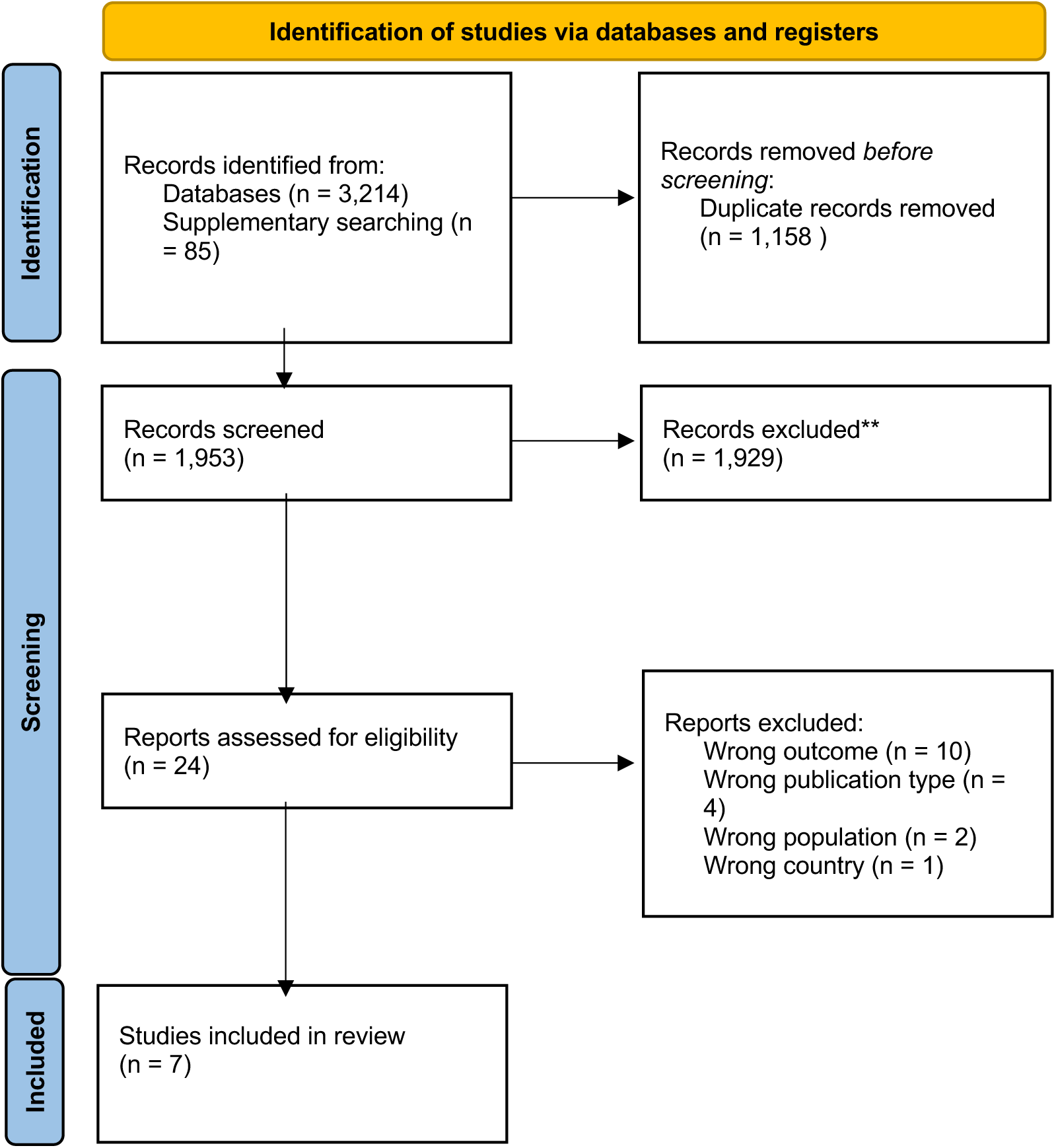
PRSIMA flow diagram.

### 6.2 Data extraction

**Table 4:** Summary of included studies.

| Citation (Country) | Study Details | Participants & setting | Key findings | Observations/notes |
| --- | --- | --- | --- | --- |
| Arman et al (2021) The effects of core stabilization exercise program in obese people awaiting bariatric surgery: A randomized controlled study, *Complementary Therapies in Clinical Practice*; 43, p.101342.<br>Turkey | **Study Design:**<br>Randomised controlled trial<br>**Study aim:**<br>The aim of this study was to compare the effects of the core stabilisation exercise program (CSEP) and physical activity counselling on functional capacity, physical fitness, physical activity, fatigue and quality of life (QoL) in obese people awaiting bariatric surgery.<br>**Type of intervention [exposure]:**<br>A twice weekly, eight-week Core Stabilisation Exercise Program consisting of a combination of strengthening, endurance and balance exercises accompanied by breathing.<br>**Comparator intervention or control:**<br>All participants received physical activity counselling.<br>**Data collection methods and dates:**<br>Participants were recruited between October 2019 and March 2020. Assessments were conducted at the start and end of the study (after eight weeks).<br>**Outcomes (relevant):**<br>Functional capacity (6-minute walk test)<br>Body composition (bioimpedance analysis) | **Sample size:**<br>23 (21 completed)<br>Intervention Group: n= 10,<br>Control Group: n= 11<br>**Participants:**<br>Adult candidates with morbid obesity (BMI $\geq 40$) awaiting bariatric surgery were eligible if they were expected to be operated on within 3–6 months. Participants were referred to the research project by their surgical teams. Exclusion criteria for candidates were as follows; unable to regularly attend supervised exercise sessions from 6 months, have musculoskeletal disorders or systemic diseases that may prevent exercise, presence of uncontrolled neuropsychiatric illnesses affecting cooperation and cognitive functions, presence of acute pain or heart pain and previous myocardial infarction, subjective heart failure, uncontrolled diabetes and hypertension.<br>**Setting:**<br>Department of Physiotherapy and Rehabilitation, Faculty of Health Science, Istanbul University-Cerrahpasa | **Functional capacity outcomes:**<br>The intervention group significantly improved their 6 MWT-distance values after eight weeks (IG: mean difference = $146 \pm 42.33$ [CI: 115.71, 176.28]). The control group 6 MWT-distance values significantly worsened after eight-weeks (CG: mean difference = $-36.63 \pm 50.91$ [CI: -70.83, -2.43]). There was a statistically significant difference in 6 MWT-distance scores between the groups after the program in favour of the exercise group ($P < 0.05$). Cohen's d indicated on all scores of 6 MWT ranged from 0.03 to - 0.78 for the exercise group and - 0.05 to 0.26 for the control group.<br>**Body composition outcomes:**<br>The changes in all body composition scores (body weight, BMI, fat mass, fat free mass) were not statistically significant in both groups after the programs ($p > 0.05$). No between-group differences occurred for any body composition outcomes ($p > 0.05$).<br>**Fatigue outcomes:**<br>Significant improvements in FSS and FACIT-F was found in the only exercise group (FSS score: effect size = 0.79), no significant change | Of 86 patients assessed for eligibility, 56 were considered eligible. Of these, 23 agreed to participate and were randomised. Of which one patient dropped out of the study due to a lack of adherence to the program and the continuation of sedentary behavior. One patient failed to come to the clinic due to a transportation problem. |
|  | Quality of life (Obesity Specific Quality of Life questionnaire [OSQOL])<br>Fatigue (Fatigue Severity Scale and the Functional Assessment of Chronic Illness Therapy-fatigue Scale) |  | occurred in the control group. Moreover, the difference between pre-and post-tests was statistically significant between the exercise and control groups and ($p < 0.01$). The effect sizes were 0.79 and 0.59 for the scales, respectively and moderate for both scales.<br>**Quality of life outcomes:**<br>OSQOL scores were statistically significant in both groups ($p < 0.01$). The intervention group OSQOL score significantly improved whereas the control groups worsened. The effect sizes were 0.41 and -0.48 for the intervention group and control group respectively. |  |
| Baillot et al (2024)<br>Acceptability and Feasibility of the Telehealth Bariatric Behavioural Intervention to Increase Physical Activity before Bariatric Surgery: A Single-Case Experimental Study (Part I), *Obesity Surgery*, 34(5), pp.1639-1652.<br>Canada | **Study Design:**<br>Quasi-experimental before and after<br>**Study aim:**<br>1) assess the feasibility and acceptability of the TELEhealth BARIatric behavioural intervention (TELE-BariACTIV) trial protocol/methods and intervention, which was designed to increase moderate-to-vigorous intensity physical activity (MVPA) in adults awaiting MBS; and 2) estimate the effect of the intervention on MVPA.<br>**Type of intervention [exposure]:**<br>Six weekly 45-minute PA counselling sessions via videoconferencing designed to increase MVPA.<br>Participants were randomly assigned to either a 1- or 2-week baseline A1 phase; subsequent phases (phases B1 and A2) were of identical length for all participants. The A phases were observational phases | **Sample size:**<br>12<br>**Participants:**<br>Aged $\geq 18$ years; ii) scheduled to undergo a sleeve gastrectomy $\geq 12$ weeks at one of the three participating hospitals; iii) self-reported $\leq 150$ minutes of MVPA per week, and iv) had access to a computer with Internet and an interface with a camera. Exclusion criteria were: i) having a physical contraindication to physical activity; ii) already enrolled in a supervised exercise intervention or physical activity behavioural change intervention; iii) unable to speak and understand French, or iv) needing a wheelchair, cane, walker, or other assistive device(s) to move. | **Psychosocial and quality of life outcomes:**<br>At least one minimal clinically important difference for psychosocial outcomes (anxiety and depressive symptoms, quality of life) was reached by 10/11 (90.9%) participants during phase B1 (during the intervention) and during phase A2 (post-intervention) | The outcomes included in this data extraction were secondary outcomes designed to assess the feasibility of a larger-scale intervention in the future and may lack appropriate statistical analysis.<br>Intervention acceptability and engagement outcomes were reported by the study; however, they were not comparative and therefore not extracted by the review team. |
|  | without intervention, and the B phases were interventional with the TELE-BariACTIV. Daily MVPA was assessed for 7 (if randomised to 1 week baseline A1) or 14 days (if randomized to 2-week baseline A1) during the A1 phase, and then for 7 days in phases B1 and A2.<br>**Data collection methods and dates:**<br>Accelerometer, validated questionnaires, and self-reported measures of data collection. Recruitment took place between September 2021 and July 2022.<br>**Outcomes (relevant):**<br>• Anxiety (GAD-7)<br>• Depressive symptoms (PHQ-9)<br>• Quality of life (RAND) | **Setting:**<br>The intervention was delivered remotely. Participants were recruited from three hospitals in Canada, two associated tertiary care centres and one associated regional centre. |  |  |
| Baillot et al (2017)<br>Feasibility and effect of in-home physical exercise training delivered via telehealth before bariatric surgery, *Journal of telemedicine and telecare*, 23(5), pp.529-535.<br>Canada | **Study Design:**<br>Quasi-experimental (with historical comparison groups)<br>**Study aim:**<br>To assess the feasibility and effect of Pre-Surgical Exercise Training (PreSET) delivered in-home via telehealth (TelePreSET) in subjects awaiting bariatric surgery.<br>**Type of intervention**<br>**[exposure]:** Twelve-weeks of endurance and strength training, supervised twice weekly using videoconferencing.<br>**Comparator intervention or control:**<br>Pre-post as well as a usual care group from a previous study (Baillot et al 2016).<br>**Data collection methods and dates:**<br>Satisfaction and weight-related quality of life were assessed at baseline and after 12-weeks (post-intervention) using validated | **Sample size:**<br>6<br>**Participants:**<br>Women awaiting bariatric surgery (aged ≥ 18 years; body mass index (BMI) ≥ 35 with comorbidities or ≥ 40 kg/m<sup>2</sup>), who were expected to be operated within 3–6 months. Exclusion criteria included: medical contraindication for physical activity, functional limitations, >one weekly supervised exercise sessions, no access to high speed residential Internet, and not understanding French.<br>**Setting:**<br>Participants homes. | **Satisfaction outcomes:**<br>The baseline in-home telehealth patients' perception score was high (83.5% (80.9–91.2), and increased significantly after the intervention (90.0% (86.8–94.1); p=0.03). The global TelePreSET' satisfaction was 93.4% (89.3–97.7), with subscores of 97.5% (91.9–98.1) for quality of care provided, 90.0% (78.0–97.0) for similarity to face-to-face encounter and 100.0 % (95.0–100.0) for perception of the interaction.<br>**Weight-related quality of life outcomes:**<br>No difference was also found between the TelePreSET and the usual care groups (p≥0.4), with a trend for difference in the confidence during exercise (7.5 (2.5–10.0) vs 0.0 (-15.0–5.0); p=0.06) and social interaction | Comparison groups were used from Baillot et al (2016) study which compared a supervised in-person exercise intervention (PreSET) to usual care.<br>Five women for bariatric surgery refused to be contacted, and 21 were contacted by telephone between March–October 2014. Among the 21 women, 12 refused to participate (no time for exercise training, n=7; moving, n=2; not interested by research or telehealth, n=3), and nine were excluded due to insufficient space for training (n=5), functional limitation (n=1), current involvement in supervised exercise (n=2) and inability to understand French (n=1). |
|  | questionnaires. Participants recruited between March-October 2014.<br>**Outcomes (relevant):**<br>• Satisfaction with telehealth<br>• Weight-related quality of life (The Laval questionnaire)<br>• Anthropometric outcomes: BMI, fat mass. |  | scores (11.2 (1.0-28.6) vs 2.0 (-10.2-2.0); $p=0.05$).<br>**Anthropometric outcomes:**<br>No significant change was found in anthropometric, body composition and blood pressure after 12 weeks in each group, and no significant differences between groups were observed ($p>0.2$). |  |
| Kulinski and Smith (2020) Surgical prehabilitation using mobile health coaching in patients with obesity: A pilot study, *Anaesthesia and Intensive Care*, 48(5), pp.373–380<br>Australia | **Study Design:**<br>Quasi-experimental before and after pilot study.<br>**Study aim:**<br>To test the implementation of a mobile phone-based text message prehabilitation programme for patients with obesity awaiting elective surgery.<br>**Type of intervention [exposure]:**<br>Mobile phone-based automated text message prehabilitation programme. A bank of 96 messages from the fields of nutrition, exercise, psychology and medical management, based on Australian guidelines. Messages were sent four times per week for six months. Links to local health resources also provided, e.g. parkrun, local council activities.<br>**Comparator intervention or control:**<br>Outcomes compared to baseline.<br>**Data collection methods and dates:**<br>Participant-reported including physical and behavioural measures, subjective experience, and a validated scale. Feasibility also assessed. Participants recruited between October 2018 and May 2019. | **Sample size:**<br>22 (18 completed the study)<br>**Participants:**<br>Adults with a BMI of 30 and above, listed for a procedure with a 12-month waiting time non-pregnant and owning a mobile phone. Exclusion criteria included: participants had to own a mobile phone, 30+ BMI.<br>**Setting:**<br>Patients were recruited from the surgical waitlist within the Illawarra-Shoalhaven local health district. | **Weight:**<br>Eight out of 18 participants (40%) lost at least 2 kg by the conclusion of the programme. Those who did lose weight lost a mean of 6 kg over six months (range 2– 12 kg), with a mean weight loss of 1.7 kg for the entire cohort. 8 (40%) reported improved BMI change. Participant 5 experienced a 12 kg weight loss and reported that they cancelled their orthopaedic procedure as a result.<br>**Health-related quality of life:**<br>Eleven participants (61%) reported that their overall 'health score' as indicated by the EuroQol visual analogue scale improved after completing the programme. 5 of 18 reported improved general health-related quality of life. | 13/75 refused enrolment. Of the four non-completing patients, one was unable to participate by the end of the study due to a new health diagnosis, and three were unable to be contacted at the completion of the programme.<br>2/18 participants thought that 4 messages a week were too many. |
|  | **Outcomes (relevant):**<br>Weight<br>Economics (generic health quality of life measure, EQ-5D-3L) |  |  |  |
| Lau et al (2024)<br>Psychological Benefits of a Preoperative Educational Bridging Program for Bariatric Surgery: Does Face-to-Face versus Videoconference-based delivery make a difference?, *Obesity Facts*<br>Germany | **Study Design:**<br>Quasi-experimental before and after<br>**Study aim:**<br>To assess the effects of a group-based educational programme before MBS on three key factors: i) patients' mental health, ii) the program's perceived helpfulness from the patients' perspective, and iii) the effectiveness of delivering the program online via videoconferencing.<br>**Type of intervention [exposure]:**<br>Preoperative Educational Bridging Programme. The programme lasts three months and consists of four theoretical and three exercise sessions.<br>Originally delivered in-person, but conducted online via video conferencing since the COVID-19 pandemic. The face-to-face sessions were attended by 12 participants and each session lasted 90 minutes, while the videoconference-based sessions had no more than 10 participants and lasted one hour each.<br>**Comparator intervention or control:**<br>Baseline, comparisons also analysed between face-to-face and online.<br>**Data collection methods and dates:**<br>Validated questionnaires, data collected and analysed between May 2014 and August 2022.<br>**Outcomes (relevant):** | **Sample size:**<br>305<br>Face-to-face group: n = 222, online group: n = 66, both formats: n = 17<br>**Participants:**<br>Adults with a BMI over 40, or greater than 35 with comorbidities awaiting bariatric surgery. These patients have i) undergone conservative therapies according to the guidelines but still need to exhaust conservative methods before qualifying for MBS as the next-step in guideline-based therapy or ii) have a primary indication for surgery, such as severe weight-related diseases that preclude conservative treatment.<br>**Setting:**<br>Face-to-face group: University Hospital Tuebingen.<br>Online group: participants homes | **Total group:**<br>**Anxiety outcomes:**<br>Anxiety symptoms significantly improved during the intervention, with a mean reduction of 1.1 units (SD 4.6, $z = -3.914$, $p < 0.001$).<br>**Depression outcomes:**<br>Depression symptoms significantly improved, with a mean reduction of 0.9 units (SD 4.6, $z = -3.771$, $p < 0.001$.)<br>**Stress outcomes:**<br>Overall stress reduced between T0 and T1 by 4.6 (SD 15.6) points ($z = -4.976$, $p < 0.001$). In particular, the subscale "worries" showed an average improvement of -7.9 points (SD 22.7, $z = -5.761$, $p < 0.001$). A mean reduction of 5 points was achieved for the subscale "tension" ($z = -4.237$, $p < 0.001$) and for the subscale "demands" ($z = -4.944$, $p < 0.001$). There were no significant changes on the "joy" subscale.<br>**Quality of life outcomes:**<br>There was no significant change in the physical component between T0 and T1. However, there was a trend toward improvement in the mental component of quality of life, with a mean increase of 2.3 points (SD 10.8, $z = 3.022$, $p = 0.003$).<br>**Weight & BMI outcomes:** | The drop-out rate was 2.8% (n = 10 participants). There were no significant differences between completers and dropouts at baseline. |
|  | • Anxiety (GAD-7)<br>• Depression (PHQ-9)<br>• Stress (PSQ-20)<br>• Quality of life (SF-12)<br>• Weight<br>• BMI<br>• Satisfaction |  | There were almost no changes in weight (-0.3 kg (SD 8.7, $z = -0.561$, $p = 0.575$) and BMI (-0.1 units, SD 3.1, $z = -0.277$, $p = 0.782$) during the intervention.<br>**Face-to-face and videoconference-based**<br>Both the face-to-face and videoconference subgroups showed improvements over time in anxiety, mental quality of life and for perceived stress on the overall stress scale and on the three subscales “worries”, “tension”, and “demands”. Depressive symptoms and quality of life remained unchanged from T0 to T1. There were no interaction effects between the two subgroups indicating that the mode of delivery (face-to-face vs. videoconference) did not significantly influence outcomes. In addition, there were no statistically significant differences between the MCIDs achieved by the two subgroups, supporting the similar effectiveness of the two delivery modes. |  |
| Marc-Hernández et al (2019) Impact of Exercise on Body Composition and Cardiometabolic Risk Factors in Patients Awaiting Bariatric Surgery. *Obesity Surgery*, 29, pp.3891-3900<br>Spain | **Study Design:**<br>Non-randomised controlled study. (Patients were assigned to the experimental or control group in chronological order of recruitment: the first 12 patients were included in the exercise group (EG) and the other 11 patients in the control group).<br>**Study aim:**<br>To describe the effects of an exercise programme on body composition and cardiometabolic risk factors in patients awaiting bariatric surgery. | **Sample size:**<br>23<br>Intervention group: $n = 12$, control group: $n = 11$.<br>**Participants:**<br>Adults with a BMI of 40 or above or 35 with an obesity-associated comorbidity, awaiting bariatric surgery. Exclusion criteria were as follows: (a) cardiovascular diseases; (b) severe functional limitations; (c) chronic respiratory diseases; or (d) plans to undergo | **Body composition (bioimpedance) & BMI outcomes:**<br>The exercise group (EG) obtained significant reductions in all anthropometric and body composition variables evaluated except in the FFM, which remained constant; total weight ($-7.3 \pm 4.1$ kg, $p < 0.01$), fat mass ($-7.1 \pm 4.7$ kg, $p < 0.01$), and waist circumference ($-5.3 \pm 2.1$ cm, $p < 0.01$). All these variables, excluding FFM, showed significant differences between groups. | Two women of the intervention group did not finish the exercise program (one left the program and another experienced physical problems). |
| | **Type of intervention [exposure]:**<br>A 12-week exercise programme which combined endurance and resistance training.<br>**Comparator intervention or control:**<br>Control group which received advice to follow an active lifestyle.<br>Both groups received the usual care prior to surgery, including psychological and nutritional counselling.<br>**Data collection methods and dates:**<br>Physical and biochemical tests, validated questionnaire, anthropometrics. Dates of data collection not stated.<br>**Outcomes (relevant):**<br>• Body composition (bioimpedance)<br>• Waist and hip circumference (International Society for the Advancement of Kinanthropometry protocol)<br>• BMI<br>• Excess weight loss<br>• Basal metabolic rate<br>• Health-related quality of life (SF-36) | another exercise program during the period of the study.<br>**Setting:**<br>Not stated. | The EG had significantly greater reductions than the control group (CG) in the percentage of BMI reduction ($-5.6 \pm 3.32\%$ vs $-0.57 \pm 1.95\%$, $p = 0.002$), fat mass ($-10.93 \pm 7.73$ vs $-1.75 \pm 3.05\%$, $p = 0.005$), and visceral fat ($-11.28 \pm 7.49\%$ vs $-2.33 \pm 5.20\%$, $p = 0.011$).<br>**Waist and hip circumference outcomes:**<br>Waist-to-height ratio only decreased significantly in the EG ($p = 0.007$).<br>**Excess weight loss (EWL) outcomes:**<br>The %EWL was of $15.2 \pm 11.2\%$ vs. $1.5 \pm 5.2\%$ in the EG and the CG, respectively ($p = 0.006$; 95% CI, 4.6–22.8; Student's t test).<br>**Basal metabolic rate outcomes:**<br>Significant changes in the basal metabolic rate were not observed in any of the groups.<br>**Health-related quality of life outcomes:**<br>the EG increased their scores on all scales of the SF-36. These increases were significant in 3 of the 8 scales of the questionnaire (physical functioning, $p = 0.002$, 95% CI, 11.7, 39.2; general health perceptions, $p = 0.003$, 95% CI, 8.0, 27.6; mental health, $p = 0.021$, 95% CI, 1.9, 18.8), and in one of the scales, social functioning, there was a large increase ($d = 0.81$, 95%CI, $-0.13, 1.68$). In addition, the EG also showed a significant<br>increase in the physical component summary ( $p = 0.004$ , 95% CI, 3.1, 12.3). The CG showed significant increases in physical role ( $p = 0.042$ , 95% CI, 1.1, 40.6), a small change in bodily pain ( $d = 0.43$ , 95%CI – 0.75, 1.54) and a moderate change in emotional role ( $d = 0.52$ , 95% CI – 0.67, 1.63). A significant difference between groups was obtained in absolute changes in physical functioning, general health perceptions, and physical component summary. | |
| Tokgoz et al (2022) Effects of pre-surgical aerobic dance-based exercise on lower extremity in people with morbid obesity awaiting bariatric surgery: Randomized controlled study, *Clinical Obesity*, 12(4), pp.12529.<br>Turkey | **Study Design:**<br>Randomised controlled trial.<br>**Study aim:**<br>To investigate what are the effects of pre-surgical aerobic dance-based exercise programme (PSADBE) and physical activity counselling (PAC) programme on lower extremity functions after surgery in people with morbid obesity awaiting bariatric surgery (PMOABS).<br>**Type of intervention [exposure]:**<br>Pre-surgical aerobic dance-based exercise for 60-minutes twice-weekly for 8-weeks, under physiotherapist supervision in groups. Both groups received PAC.<br>**Comparator intervention or control:**<br>Physical activity counselling. Individually set for each participant, included a brochure explaining importance of physical activity, examples and recommendations, participants recorded step counts and activity levels for 8-weeks.<br>**Data collection methods and dates:** | **Sample size:**<br>34 (4 dropped out)<br>Intervention group: $n=13$, control group: $n=17$.<br>**Participants:**<br>Adults with morbid obesity scheduled for laparoscopic sleeve gastrectomy within 3–6 months who had not undergone any surgical operation at the last 6 months. The exclusion criteria were: any musculoskeletal disorder, systemic illness, balance problem, psychiatric or neurological disease, acute pain or heart pain for any reason, history of myocardial infarction within 1 year, congestive heart failure, and inability to adapt to treatment.<br>**Setting:**<br>Not stated, but aerobic dance-based exercise programme was delivered in-person in groups. | **Quality of life outcomes:**<br>A statistically significant improvement was observed in all comparisons of IWQOL-LITE Total and IWQOL-LITE Physical Function Scores in intervention group. It was statistically significant only for comparisons of T1–T2 and T1–T3 in the control group ($p < .05$). Intergroup comparisons found statistically significant differences in favour of the intervention group at T1-T2 for both IWQOL-Lite total (mean difference: IG= $-14.46 \pm 16.46$, CG= $1.64 \pm 22.08$, $p = 0.007$) and IWQOL-LITE Physical Function Scores mean difference: IG= $-10.23 \pm 6.35$, CG= $1.70 \pm 8.01$, $p = 0.001$). No statistically significant between-group differences were apparent at T1-T3.<br>**Body composition outcomes:**<br>A statistically significant improvement was found in all parameters for comparisons of T1–T3 and T2–T3 in both groups ($p < .05$). | Fifty-six patients were evaluated for eligibility for the study. Thirty-four patients were included in the study. During the study, 4 patients dropped out (personal reasons $n=2$, transfer problems $n=2$). |
|  | Physical exercise tests, bioimpedance analysis, validated questionnaires and scales. Groups were evaluated before exercise (T1), after an 8-week exercise programme (T2), and at 5 months postoperative (T3). Data collection dates not stated but participants recruited between November 2019 and June 2020.<br>**Outcomes (relevant):**<br>• Quality of life (IWQOL-LITE)<br>• Body composition (Bioimpedance)<br>• Fatigue (Fatigue Severity Scale (FSS)) |  | The differences of all scores for between-group comparisons of T1–T2 and T2–T3 were not statistically significant ($p > .05$).<br>**Fatigue outcomes:**<br>In intergroup comparison, the improvements in the FSS scores for comparisons of T1–T2 and T1–T3 in the intervention group and the improvements in the FSS score only for comparisons of T1–T3 in the control group were found statistically significant ($p < .05$). In intergroup comparison, the differences in FSS score for comparisons of T1–T2 and T1–T3 were significant in favour of the intervention group ($p < .05$). |  |

### 6.3 Quality appraisal

**Table 5:**
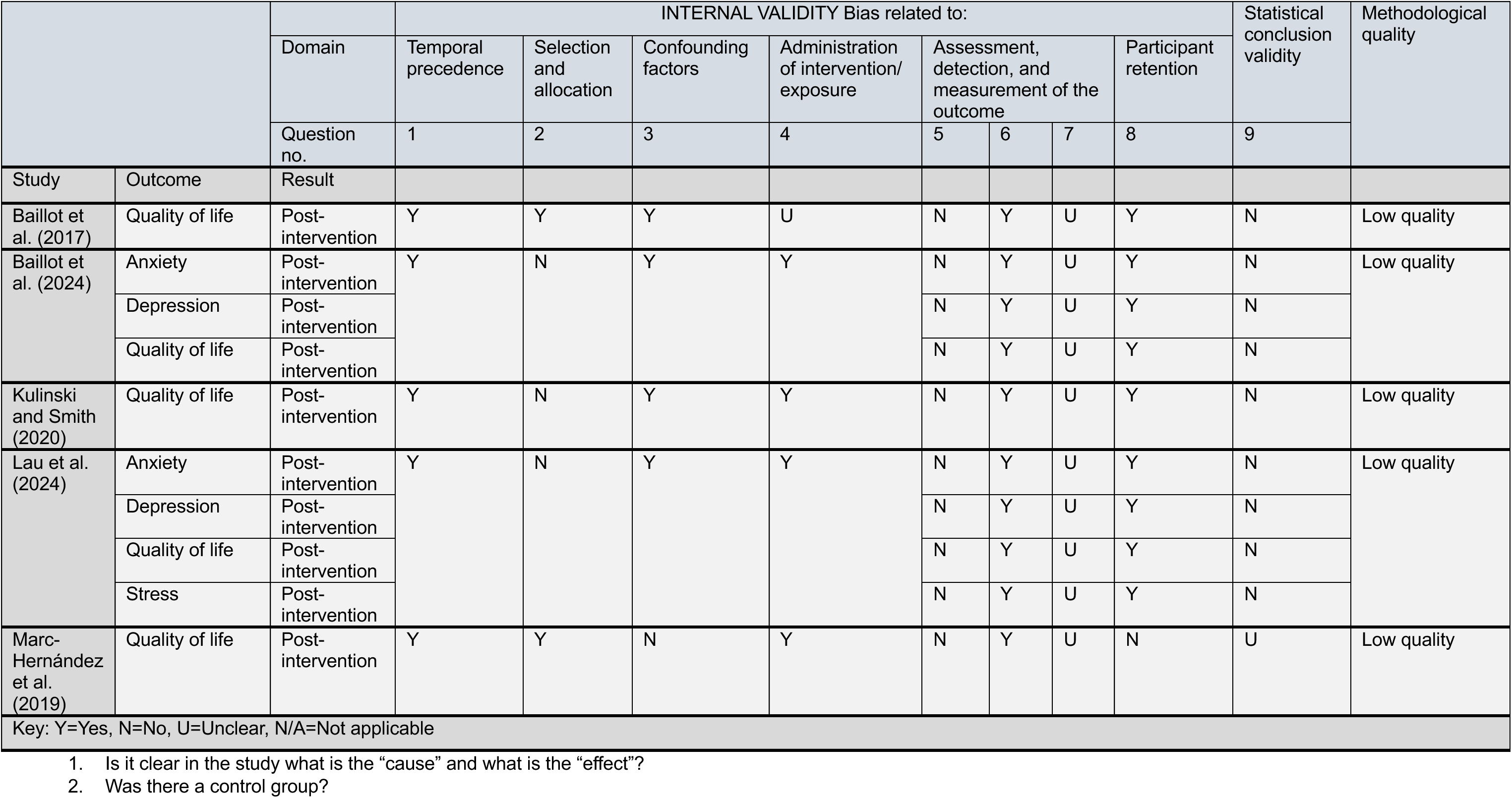
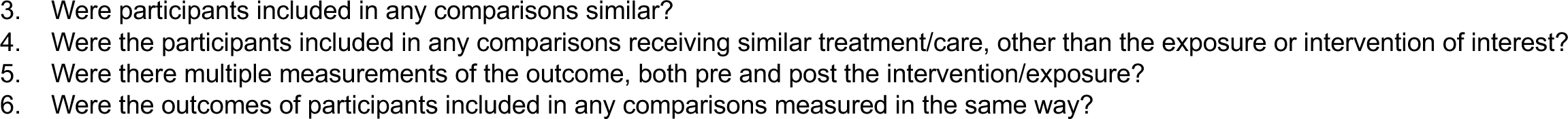
Quality appraisal results for quasi-experimental studies.

|  |  | Domain | INTERNAL VALIDITY Bias related to: |  |  |  |  |  |  |  | Statistical conclusion validity | Methodological quality |
| --- | --- | --- | --- | --- | --- | --- | --- | --- | --- | --- | --- | --- |
|  |  |  | Temporal precedence | Selection and allocation | Confounding factors | Administration of intervention/ exposure | Assessment, detection, and measurement of the outcome |  |  | Participant retention |  |  |
|  |  |  | 1 | 2 | 3 | 4 | 5 | 6 | 7 | 8 | 9 |  |
| Study | Outcome | Result |  |  |  |  |  |  |  |  |  |  |
| Baillot et al. (2017) | Quality of life | Post-intervention | Y | Y | Y | U | N | Y | U | Y | N | Low quality |
| Baillot et al. (2024) | Anxiety | Post-intervention | Y | N | Y | Y | N | Y | U | Y | N | Low quality |
|  | Depression | Post-intervention |  |  |  |  | N | Y | U | Y | N |  |
|  | Quality of life | Post-intervention |  |  |  |  | N | Y | U | Y | N |  |
| Kulinski and Smith (2020) | Quality of life | Post-intervention | Y | N | Y | Y | N | Y | U | Y | N | Low quality |
| Lau et al. (2024) | Anxiety | Post-intervention | Y | N | Y | Y | N | Y | U | Y | N | Low quality |
|  | Depression | Post-intervention |  |  |  |  | N | Y | U | Y | N |  |
|  | Quality of life | Post-intervention |  |  |  |  | N | Y | U | Y | N |  |
|  | Stress | Post-intervention |  |  |  |  | N | Y | U | Y | N |  |
| Marc-Hernández et al. (2019) | Quality of life | Post-intervention | Y | Y | N | Y | N | Y | U | N | U | Low quality |
| Key: Y=Yes, N=No, U=Unclear, N/A=Not applicable |  |  |  |  |  |  |  |  |  |  |  |  |

1. Is it clear in the study what is the “cause” and what is the “effect”? 2. Was there a control group?
3. Were participants included in any comparisons similar? 4. Were the participants included in any comparisons receiving similar treatment/care, other than the exposure or intervention of interest? 5. Were there multiple measurements of the outcome, both pre and post the intervention/exposure? 6. Were the outcomes of participants included in any comparisons measured in the same way?

**Table 6:** Quality appraisal results for randomised controlled trials.

|  |  |  | INTERNAL VALIDITY Bias related to: |  |  |  |  |  |  |  |  |  |  |  |  |  |
| --- | --- | --- | --- | --- | --- | --- | --- | --- | --- | --- | --- | --- | --- | --- | --- | --- |
|  |  | Domain | Selection and allocation |  |  | Administration of intervention/ exposure |  |  | Assessment, detection, and measurement of the outcome |  |  | Participant retention | Statistical conclusion validity |  |  | Methodological quality |
|  |  | Question no. | 1 | 2 | 3 | 4 | 5 | 6 | 7 | 8 | 9 | 10 | 11 | 12 | 13 |  |
| Study | Outcome | Result at follow up |  |  |  |  |  |  |  |  |  |  |  |  |  |  |
| Arman et al. (2021) | Quality of life | Post-intervention | Y | U | Y | N | N | Y | N | Y | U | Y | U | Y | Y | Moderate quality |
| Tokgoz et al. (2022) | Quality of life | Post-intervention | Y | N | N | N | N | Y | Y | Y | N | N | N | N | Y | Low quality |
| Key: Y=Yes, N=No, U=Unclear, N/A=Not applicable |  |  |  |  |  |  |  |  |  |  |  |  |  |  |  |  |

1. Was true randomisation used for assignment of participants to treatment groups? 2. Was allocation to treatment groups concealed? 3. Were treatment groups similar at baseline? 4. Were participants blind to treatment assignment? 5. Were those delivering the treatment blind to treatment assignment? 6. Were treatment groups treated identically other than the intervention of interest? 7. Were outcome assessors blind to treatment assignment? 8. Were outcomes measured in the same way for treatment groups? 9. Were outcomes measured in a reliable way? 10. Was follow up complete and if not, were differences between groups in terms of their follow up adequately described and analysed? 11. Were participants analysed in the groups to which they were randomised? 12. Was appropriate statistical analysis used? 13. Was the trial design appropriate and deviations from the standard RCT design accounted for in the conduct and analysis of the trial?

### 6.4 Information available on request

The following are available on request: protocol; search strategies.

## ADDITIONAL INFORMATION

### 7.2 Acknowledgements

The Public Health Wales team would like to thank Enzo Battista-Dowds, Maria Cole, Megan Elliot, Praveena Pemmasani and Libby Humphries for their time, expertise and contributions during stakeholder meetings, in guiding the focus of the review and interpretation of the findings.

## APPENDIX

### APPENDIX 1

Ovid MEDLINE(R) ALL <1946 to July 17, 2024>

1. (obese or obesity).ti,ab. 394914
2. Obesity/ or Obesity, Abdominal/ or Obesity, Morbid/ 255657
3. 1 or 2 449482
4. (awaiting or waitlist* or “wait* list*” or backlog* or (wait* adj3 (support* or interven* or assist* or program* or service* or treat* or surg* or refer* or specialist* or consult* or clinic* or care or appointment* or procedure* or therapy or therapeutic or team* or session* or (weight adj3 manag*) or (obesity adj3 manag*)))).ti,ab. 44687
5. Waiting Lists/ or Case Management/ or Patient Care Management/ or Critical Pathways/ 37247
6. “Referral and Consultation”/ and (Disease Management/ or Obesity Management/) 569
7. 4 or 5 or 6 74724
8. 3 and 7 1348
9. limit 8 to (english language and yr=“2010 -Current”) 1064
10. 9 not (case reports or comment or editorial or letter).pt. 1028

## Footnotes

* This section has been completed by the Centre for Health Economics & Medicines Evaluation (CHEME), Bangor University

